# Prevalence of anemia, associated risk factors and outcome in CHUK, Rwanda: a prospective observational study

**DOI:** 10.1101/2023.03.09.23286999

**Authors:** Phocas Havugimana, Raphael Ndahimana, Felix Babane, Ernestine Umutesi, Polyphile Ntihinyurwa, Diane Mushimiyimana, Florence Masaisa, Etienne Ntabanganyimana

## Abstract

**Background:** Admitted patients with anemia are at increased morbidity and mortality risk as well as length of hospital stay. It affects more than 2 billion people worldwide and is causing significant morbidity and mortality. Its etiology is varying with many predisposing factors including nutritional deficiencies, infections, Malignancies, chronic inflammations and other chronic diseases like autoimmune diseases, chronic liver disease, and chronic kidney disease.

**Methods:** We performed a non-randomized, prospective observational study of 143 patients admitted in Internal Medicine between March and April 2021, we assessed their hemoglobin level in order to determine the prevalence of anemia. The demographic and clinical data were collected using a designed questionnaire. All patients found to have anemia were followed for outcome assessment (either discharge or died).

**Results:** The prevalence of anemia was high (52.4%) among 143 admitted patients in Internal Medicine, CHUK. Patients with HIV, cancer and chronic kidney disease had 5.84-, 4.11- and 3.79-times risk of having anemia respectively. In 75 patients who had anemia 10 patients died among them 5 patients were having severe anemia; 25 patients were 60 years old and above; 60 patients had normocytic anemia and they had an average of length of hospital stay of 20.6 days, patients with severe anemia, length of hospital stay was 28 days.

**Conclusion:** This study demonstrated a high prevalence of anemia which is associated with high mortality rate among admitted patients in CHUK. Priority should be given to the preventive medicine, optimal management of chronic disease and geriatric medicine.

## INTRODUCTION

Anemia is defined as decreased level of hemoglobin below the normal limit of less than 13g/dl and 12g/dl in men and women respectively [1,2]. WHO categorizes anemia as mild when hemoglobin is from 10 to 11.9g/dl and 10 to 12.9 g/dl in women and men respectively; moderate when hemoglobin level is from 7 to 9.9g/dl in both genders and severe when hemoglobin is less than 7g/dl in both men and women [3,4,5]. Anemia remains a big concern worldwide, affecting all genders and all age categories across all countries but lower and middle income countries with less resources, deficiency in diet and many comorbidities are more affected [2,6,7,8]. Anemia tops all known blood abnormalities worldwide with around 28.4% of the world’s population affected around the globe [6,9]. The most affected population worldwide include women, young children, and people with chronic diseases [9]. It is more prevalent among pre-school children with 47.4% and less prevalent among men with 12.7% [6,7]. It affects more than 2 billion people worldwide and is causing significant morbidity and mortality among anemic subjects [10,11,12]. Its etiology is multifactorial in most cases with many predisposing factors including nutritional deficiencies, infections, Malignancies, chronic inflammations and other chronic diseases like autoimmune diseases, chronic liver disease, and chronic kidney disease [2,10,13]. Admitted patients with concurrent anemia are at increased risk of morbidity and mortality [9]. There is evidence that co-existing anemia prolongs hospital stay, increases re-admission rates and the outcomes in hospitalized patients can be improved by managing anemia along with the primary cause of admission [2,9,14]. From the WHO data, most cases of anemia are in Africa [4]. In Ghana, it was found that anemia was the leading cause of admission and second most common contributing factor for death [6].

In Rwanda, we don’t have enough data regarding anemia in general population. Rwanda Demography and Health Survey (DHS) 2015 report, showed the prevalence of anemia of 36.5% in children and 19.2% in females [15]. This study was evaluating the prevalence of anemia among admitted patients in CHUK, associated risk factors and outcomes in terms of mortality and hospital stay.

## METHODS

### Study design and methodology

Kigali University Teaching Hospital, CHUK is a public, tertiary hospital in Rwanda, located in Kigali, Capital City of Rwanda. It is the biggest hospital among three major referral hospitals located in Kigali with 560 beds capacity. The Internal Medicine is among the departments based on type of specialties. We performed a non-randomized, prospective observational study. Between March and April, 2021; 143 patients aged ≥ 15 years old, who were admitted in Internal Medicine, their hemoglobin was assessed to determine if they had anemia or not. The anemia was classified as normocytic anemia (means with normal mean corpuscular volume 80-100 fl), microcytic anemia (means low mean corpuscular volume), macrocytic anemia (means high mean corpuscular volume), mild anemia (hemoglobin >/=10 g/dl) moderate anemia (hemoglobin 7-10g/dl) and severe anemia (hemoglobin </= 7g/dl).

### Patient and Public Involvement

All patients admitted in Internal Medicine wards, were recruited in the study. Prior to the recruitment of the study participants, by using local language (Kinyarwanda), the patients were explained the purpose of the study as well as the potential benefits and risks of participating to the study. The investigator told them that they were free to withdraw from study at any time, without having to give a reason of withdrawing and without affecting their future medical care. They had the opportunity to ask questions. By signing the Informed Consent, they knew that they authorized access to their medical records to the monitor(s) and the auditor(s), and possibly to the members of the Ethical Committees or Health Authorities, for verification of clinical study procedures and/or data. They have been given a copy of the Informed Consent Form.

## ANALYSIS

Date entry was done using Epidata version 3.1 and exported to excel and the statistical package for social science (IBM SPSS Statistics) version 28 for final data cleaning and analysis. We performed descriptive statistics determine frequency and percentages and later we did chi-square test of independence and Binary logistic regression to find out the relevant significance of association where by p-value <0.05 under confidence of 95% the predictive variable(s) was considered as significantly associated with outcome variable. No confounding variables identified in literature review fitting for multinomial logistic regression testing for adjustment.

## RESULTS

Of 143 patients; more than half (52.4%) had anemia (Figure 1); 53.1% were male, the majority (64%) reported to get less than two meals per day; among the common comorbidities, 16.1% had cancer disease followed by chronic liver disease (15.4%) and chronic kidney diseases (12.6%); and among 22 menstruating females, 11.8% had normal menstruation period (Table 1). Among the classification; 26 patients had mild anemia and among them 4 patients died; 13 patients had severe anemia and among them 5 patients died; the mortality was almost the same among anemic (10 patients) and non-anemic (9 patients) (Table 2). During the assessment of associated risk factors; chronic kidney disease (OR 3.79, p=0.025), cancers (OR 4.11, p=0.009), HIV (OR 5.84, p=0.025) and other comorbidities (OR 5.9, p=0.000) showed significant association with anemia (Table 3). In assessment of severity; the majority had normocytic anemia (60 patients) followed by microcytic anemia (11 patients) and 4 patients had macrocytic anemia (Figure 2). In assessment of age group, among anemic patients, 25 patients were aged 60 years and above, followed by 16 patients who were aged 31 to 40 years old (Figure 3). In assessment of hospital stay; the average was 20 days and the severity of anemia was associated with prolonged hospital stay, where the patients who had severe anemia spent 28 days in hospital (Figure 4). In assessment of death rate; among anemic patients, 10 patients and the severity of anemia was associated with high mortality rate (Figure 5).

**Table 1.** Baseline characteristics of the enrolled participants Source: Author’s Compilation, 2021

| Characteristics | Number(n) | Percentage (%) |
| --- | --- | --- |
| **Age (Mean+/-SD) 51.6±19.6** |  |  |
| **Gender** |  |  |
| Men | 76 | 53.1 |
| Women | 67 | 46.9 |
| **Food Security** |  |  |
| Less than 2 meals per day | 91 | 64 |
| Greater than or equal 2 meals per day | 52 | 36 |
| **Comorbidities** |  |  |
| Chronic Kidney Disease | 18 | 12.6 |
| Cancers | 23 | 16.1 |
| Chronic Liver disease | 22 | 15.4 |
| HIV/AIDS | 13 | 9.1 |
| Heart Failure | 14 | 9.8 |
| Chronic Lung disease | 7 | 4.8 |
| Peptic Ulcer disease | 6 | 4.2 |
| Other Diseases/comorbidities | 40 | 28 |
| **Menstruating Female** |  |  |
| < 7 days per month | 17 | 11.8 |
| > 7 days per month | 5 | 3.4 |

**Table 2.** Types of anemia and their prevalence with its effect on mortality (Bivariate analysis) Source: Author’s Compilation, 2021

| Severity-of-anemia | Admission Outcome,<br>N (%) |  | Logistic regression |  |  |  |
| --- | --- | --- | --- | --- | --- | --- |
|  | Survived | Died | OR | 95% CI |  | P-value |
| Mild | 22 (84.6) | 4 (15.4) | 1.192 | 0.333 | 4.268 | 0.787 |
| Moderate | 35 (97.2) | 1 (2.8) | 0.187 | 0.023 | 1.542 | 0.119 |
| Severe | 8 (61.5) | 5 (38.5) | 4.097 | 1.095 | 15.326 | 0.036** |
| No anemia | 59 (86.8) | 9 (13.2) | Ref. |  |  |  |
|  |  | Hemoglobin level |  |  |  | P-value |
|  |  | Mean | SD | Minimum | Maximum |  |
| **Admission** | Died | 11.4 | 4.1 | 4.2 | 17.0 | 0.551 |
| **Outcome** | Discharge | 12.0 | 3.3 | 5.6 | 20.0 |  |

**Table 3.** Bivariate analysis of factors associated with anemia. Source: Author’s Compilation, 2021

| Variables |  | Overall, N (%) | Anemia, N (%) |  | OR (95%CI) | P-value |
| --- | --- | --- | --- | --- | --- | --- |
|  |  |  | Yes | No |  |  |
| **Age** |  |  |  |  |  |  |
| <65 years |  | 96 (67.1) | 50 (52.1) | 46 (47.9) | 0.96(0.477-1.929) | 0.909 |
| >=65 |  | 47 (32.9) | 24 (51.1) | 23 (48.9) | 1 |  |
| **Gender** |  |  |  |  |  |  |
| Male |  | 76 (53.1) | 40 (54.0) | 36 (52.2) | 0.822 (0.55-2.08) | 0.822 |
| Female |  | 67 (46.9) | 34 (45.9) | 33 (47.8) | 1 |  |
| **Food Security** |  |  |  |  |  |  |
| <2 meals per day |  | 91 (63.6) | 44 (59.4) | 47 (68) | 1 |  |
| ≥ 2 meals per day |  | 52 (36.4) | 30 (40.5) | 22 (32) | 1.45(0.73-2.81) | 0.283 |
| **Comorbidities** |  |  |  |  |  |  |
| Chronic-kidney disease | yes | 18(12.6) | 14(18.9) | 4(5.8) | 3.79(1.18-12.2) | 0.025** |
|  | No | 125(87.4) | 60(81.1) | 65(94.2) | 1 |  |
| Heart failure | Yes | 14 (9.8) | 8 (10.8) | 6 (8.7) | 1.27 (0.671-3.874) | 0.671 |
|  | No | 129(90.2) | 66(89.2) | 63(91.3) | 1 |  |
| Cancer | yes | 23 (16.1) | 18(24.3) | 5 (7.3) | 4.11(1.43-11.80) | 0.009** |
|  | No | 120(83.9) | 56(75.7) | 64(92.7) | 1 |  |
| HIV | yes | 13 (9.1) | 11 (14.8) | 2 (2.90) | 5.84 (1.24-27.4) | 0.025** |
|  | No | 130(90.1) | 63(85.1) | 67(97.1) | 1 |  |
| PUD | yes | 6 (4.2) | 3(4.05) | 3 (4.35) | 0.9 (0.18-4.768) | 0.930 |
|  | No | 137(95.8) | 71(95.9) | 66(95.6) | 1 |  |
| Other comorbidities |  | 40 (27.9) | 32 (80) | 8 (20) | 5.9 (2.43-13.849) | 0.000** |

**Figure 1.**
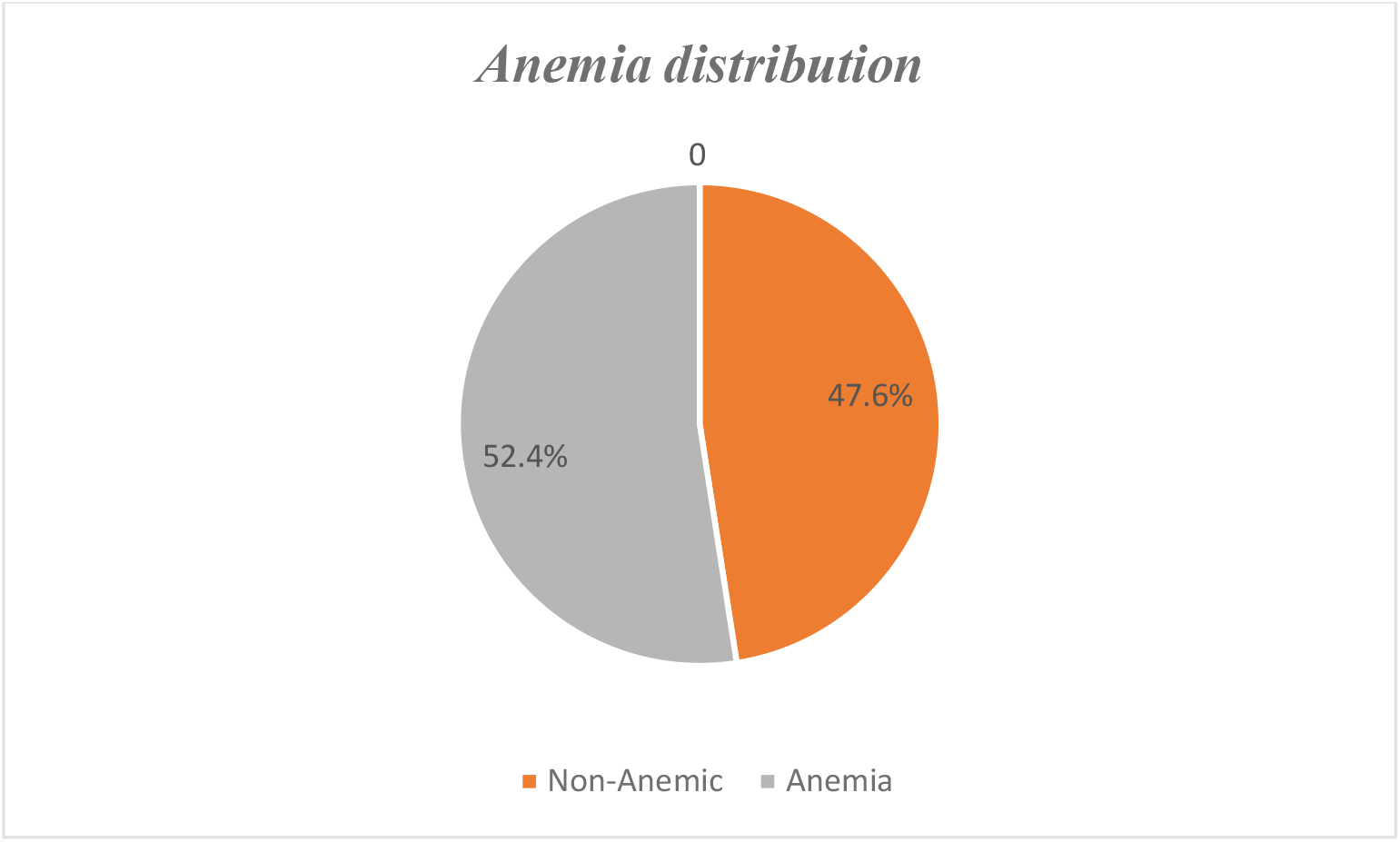
Anemia distribution. Source: Author’s Compilation, 2021

**Figure 2.**
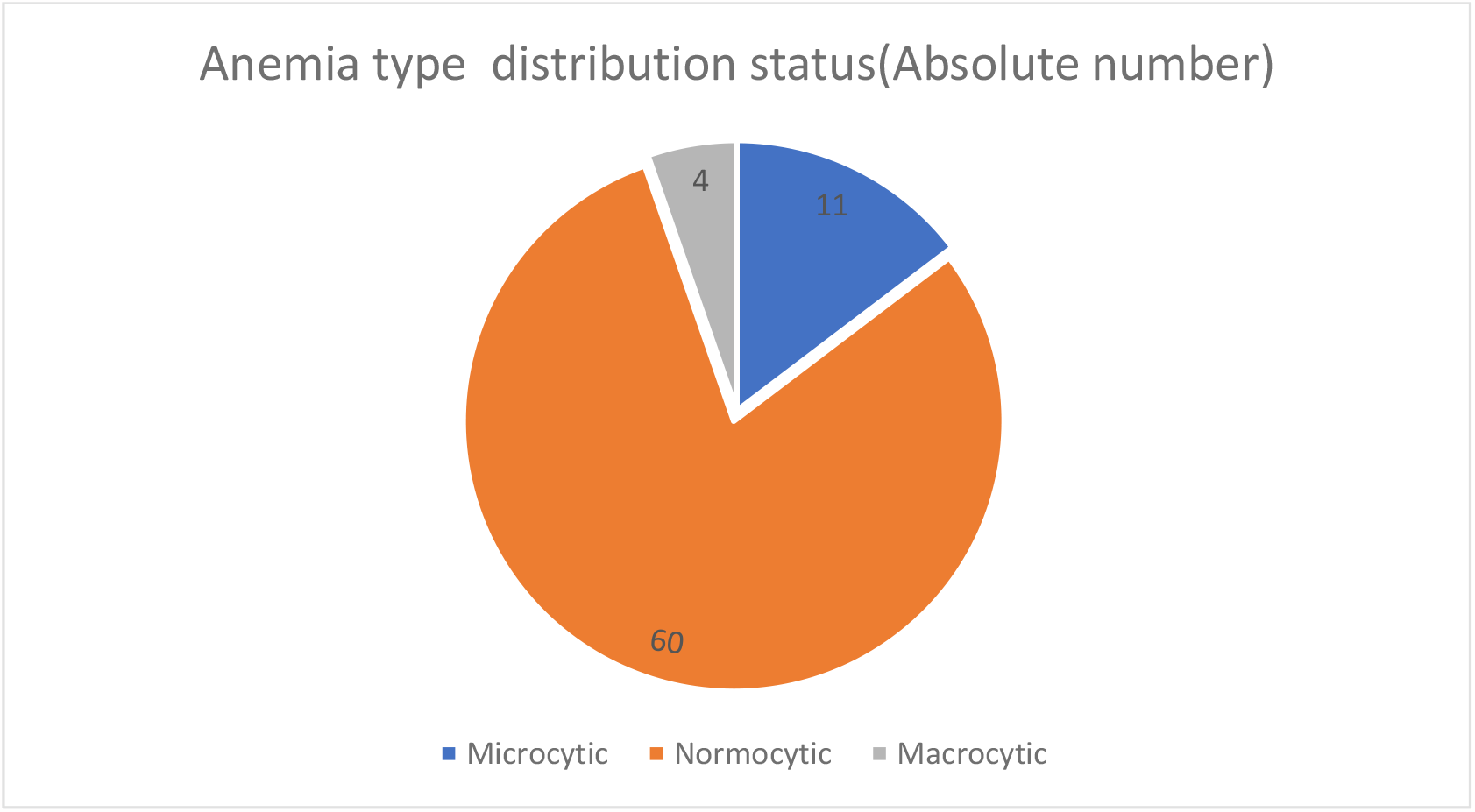
Anemia type distribution status (Absolute number) Source: Author’s Compilation, 2021

**Figure 3:**
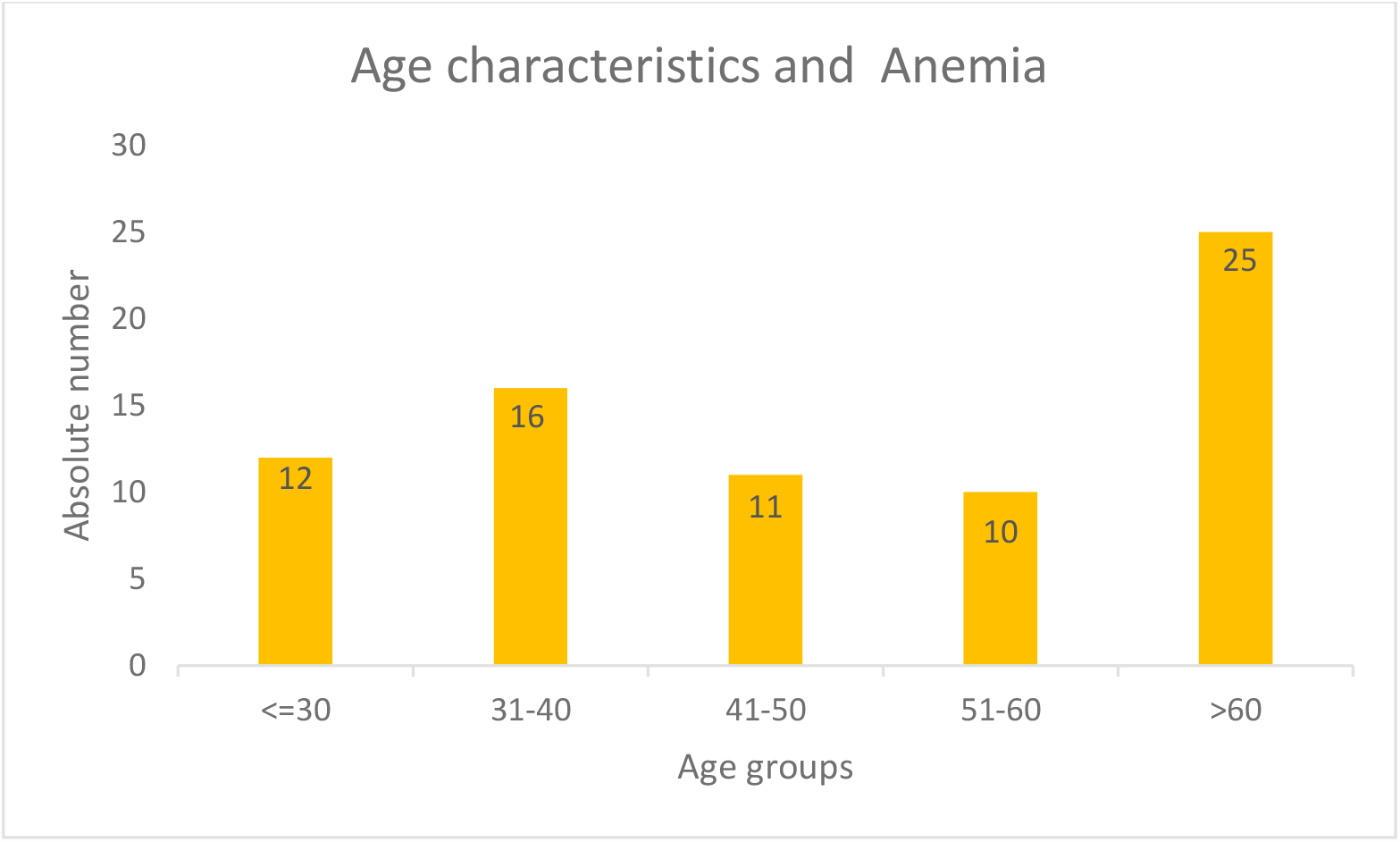
Age characteristics and Anemia Source: Author’s Compilation, 2021

**Figure 4.**
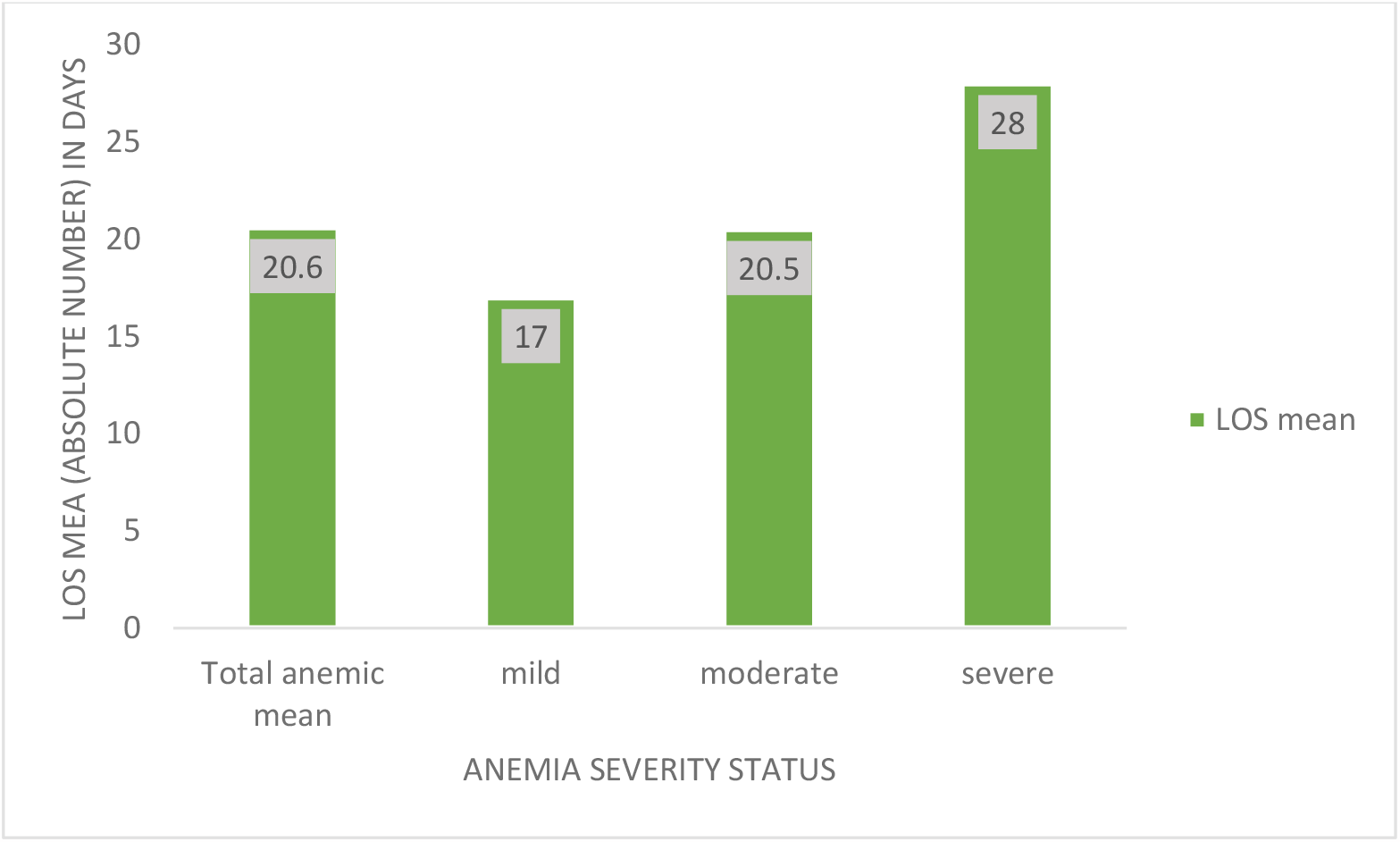
Distribution of length of hospital stay by anemia severity status. Source: Author’s Compilation, 2021 LOS: length of hospital stay

**Figure 5.**
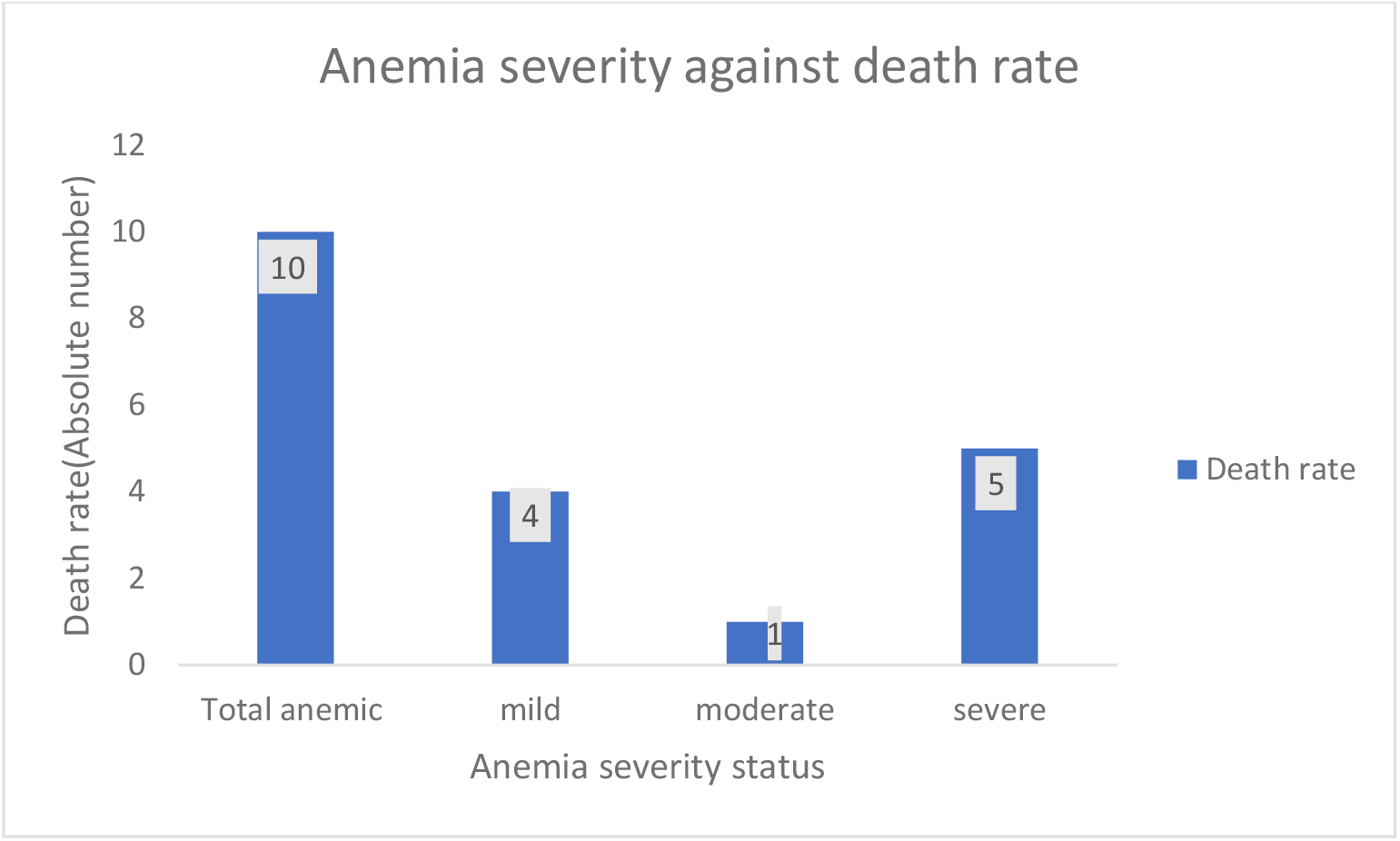
Anemia severity against death rate Source: Author’s Compilation, 2021

## DISCUSSION

Anemia is a major problem among hospitalized patients worldwide and it is associated with high mortality and morbidity [9]. The study conducted in Cooper University Hospital, US, the findings: anemic patients had overall increased mortality risk (6.5% vs. 2.5%; OR 2.68 [2.51-2.86]) than patients without anemia and anemia had an overall significant unique association with mortality (OR 4.5[3.4-6]) after adjustment for demographic factors and comorbidities [16] and another study done by Culleton BF et al. in 2006 where there was a 5-fold increase in all-cause mortality risk at Hemoglobin less than 11g/dl in unadjusted analysis (HR 5.01; 95% CI, 4.43-5.66) [17]. The study done by Zaninetti C et al, in Italy, 2017, patients with anemia had a mean length of hospital stay of 11 days, longer compared to non-anemic patients where the mean of length of hospital stay was 10 days (P-Value=0.001), the participants with severe and moderate anemia had a mean length of hospital stay of 12 days and the degree of anemia was an independent predictor of a long-term hospital stay (P-value 0.015) [18]. Another study done by Rachoin J et al supported the association of anemia and prolonged length of hospital stay with 9.8 ± 14.1 days versus 5.35 ± 8.7 days; P< 0.001) in non-anemic patients [16]. The length of hospital stay was inversely correlating with Hemoglobin level in each patient and all levels of anemia were an independent risk factor for a longer hospitalization (P=0.003, RR=1.88, CI 95% =1.3-2.85) [19].

The Prevalence of anemia in Africa is diverse. There is a reported study in Uganda with a prevalence of 16.8 to 33.8% among adults with all genders. Another study reported a prevalence of 12.5 % and 13.2% in older men and women respectively in South Africa and 23 % prevalence in Zimbabwe for general population. In Ghana a study reported as much as 53.2% prevalence in pregnant women. In Ethiopia, studies have reported the prevalence of anemia ranging from 17 to 52.3% and WHO data indicates that the prevalence of anemia among non-pregnant women in Ethiopia is around 23.3% as of 2016 [20].

Our findings in CHUK, compared to the neighboring countries, there is high prevalence of anemia among the admitted patients in Internal Medicine department where among 143 recruited patients, 52.4% had anemia (Figure 1). It is more common in senior citizens (Figure 3), normocytic anemia which known to be anemia of chronic disease [10], is more common and this is supporting the significant association between anemia and HIV, cancer, and chronic kidney disease; those mentioned diseases are chronic diseases (Figure 2, Table 3). Among 75 patients who were found to have anemia, 10 (13.3%) patients died, this is mortality rate is higher compared to the 6.5% which was found in study conducted in anemic patients in Cooper University Hospital, US and half of the death was in severe anemia (Figure 5). Our anemic patents had prolonged length of hospital stay with the mean of 20.6 days (Figure 4) which is longer than 11 days found in study conducted by Zaninetti C et al and other different studies. Also, our finding showed length of hospital stay inversely correlating with Hemoglobin level.

## CONCLUSIONS AND RECOMMENDATIONS

Based on our findings, anemia is more prevalent among the patients admitted in Internal Medicine department of CHUK. It is more common in chronic senior citizens patients and they are having prolonged length of hospital stay. At the best of our knowledge, this is the first study assessing the prevalence of anemia, associated risk factors and outcome in CHUK, as well in Rwanda. We are recommending the Ministry of Health through Rwanda Biomedical Centre, to put enough effort in preventive medicines, to optimize the management of chronic disease as the anemia is among their complications and to introduce geriatric medicine as anemia is more prevalent in people with 60 years old and above. All mentioned measures will help to control anemia and limit its morbidity and mortality.

Due to the lack of external funds, we were not able to do further deep investigations in order to get clear diagnosis.

## Data Availability

The dataset is well maintained by the study investigators and can be shared on demand

## Acknowledgements

We acknowledge all clinical staff of CHUK, especially of Internal Medicine department and all patients who accepted to participate in this study.

## Authors’ contribution

**PH** and **EN** were responsible for literature search and drafting of the article, **PH, EU, EN, RN, FB, PN** and **DM** were responsible for conception of the article, **EN** and **FM** were responsible for revision of the article.

## Ethics approval

This study was approved by the Institutional Review Board of College of Medicine and Health Sciences (N º 063/CMHS IRB/2021) and Ethics committee of CHUK (Ref: EC/CHUK/045/2021). Prior to data collection, an informed consent was obtained from the patients or the next of kin (who is legally accepted) in case the patients are not able to give his/her consent.

## Data availability

The dataset is well maintained by the study investigators and can be shared on demand

